# Longitudinal changes in physical activity during and after the first national lockdown due to the COVID-19 pandemic in England

**DOI:** 10.1101/2021.04.21.21255861

**Authors:** Feifei Bu, Jessica K Bone, John J Mitchell, Andrew Steptoe, Daisy Fancourt

## Abstract

**Background:** Recent studies have shown reduced physical activity at early stages of the COVID-19 pandemic. However, there is a lack of investigation on longitudinal changes in physical activity beyond lockdowns and stay at home orders. Moreover, it is unclear if there is heterogeneity in physical activity growth trajectories. This study aimed to explore longitudinal patterns of physical activity and factors associated with them.

**Methods:** Data were from the UCL COVID -19 Social Study. The analytical sample consisted of 35,915 adults in England who were followed up for 22 weeks from 24^th^ March to 23^rd^ August 2020. Data were analysed using growth mixture models.

**Findings:** Our analyses identified six classes of growth trajectories, including three stable classes showing little change over time (62.4% in total), two classes showing decreasing physical activity (28.6%), and one class showing increasing physical activity over time (9%). A range of factors were found to be associated the class membership of physical activity trajectories, such as age, gender, education, income, employment status, and health.

**Interpretation:** There is substantial heterogeneity in longitudinal changes in physical activity during the COVID-19 pandemic. However, a substantial proportion of our sample showed persistent physical inactivity or decreasing physical activity. Given the well-established linked between physical activity and health, persistent or increased physical inactivity is likely to have both immediate and long-term implications for people’s physical and mental health, as well as general wellbeing. More efforts are needed to promote physical activity during the pandemic and beyond.

**Funding:** Nuffield Foundation, UK Research and Innovation, Wellcome Trust

## Introduction

Since December 2019, there has been an outbreak of coronavirus disease (COVID-19). Lockdowns and ‘stay-at-home’ orders have been announced globally to control the spread of the disease, disrupting people’s usual behaviours. In many countries, this has involved the closure of gyms and outdoor sports amenities, as well as limits on how often people could leave their homes, which may have led to changes in physical activity levels. Given the beneficial impacts of physical activity for both physical and mental health,^1–3^ the negative impacts of even short-term physical inactivity,^4,5^ and the recurring nature of COVID-19 lockdown restrictions, it is important to understand how population-level physical activity has changed throughout the pandemic.

Large cross-sectional surveys comparing self-reports of activity before and after the introduction of lockdown measures have shown decreases in overall physical activity in many western countries.^6–9^ The majority of longitudinal studies to date also show an initial drop in physical activity levels following lockdown restrictions,^10–15^ as do data from wearable fitness trackers.^16–18^ However, some wearable fitness tracker studies suggest that activity levels may not have uniformly decreased across populations.^16–18^ Additionally, population-level data on the frequency of google searches for terms related to ‘exercise’ demonstrate increased interest in physical activity during lockdown (even after adjustment for increased searches on other topics).^19^ Overall physical activity may have increased following an initial drop, or the effects of lockdowns and broader social restrictions on physical activity may be more nuanced. Studies examining longer-term impacts on physical activity have provided mixed findings on whether physical activity returned to pre-lockdown levels with the initial easing of restrictions in the UK in May 2020.^10,11,14,15^

To our knowledge, no studies have investigated changes in physical activity after further easing of restrictions in the UK in July 2020, meaning the longer term impact of lockdown measures on physical activity remains unclear. We do not know whether short-term disruptions to physical activity lead to long-term behavioural changes that could have adverse effects on health. Moreover, studies to date have assumed one homogeneous trajectory of physical activity in the population, without exploring potentially heterogeneous patterns of longitudinal changes. People may have reacted differently to the pandemic and lockdown measures and therefore be at higher or lower risk of sustained changes to their physical activity levels. It is also important to identify individual characteristics that may influence physical activity. Being female, younger, single, a parent, and from an ethnic minority group as well as having poor health, lower education and income, and no access to outside space have been associated with lower physical activity during lockdown.^11,12,15,20,21^ However, findings have been inconsistent, and many studies have only examined physical activity at one point early in the pandemic. It therefore remains unclear whether similar factors are associated with trajectories of physical activity throughout lockdown and the easing of restrictions.

This study aimed to examine the heterogeneity in the longitudinal changes in physical activity in England during the initial strict lockdown and the following easing of restrictions, using a sample of 35,915 adults tracked for 22 weeks (24^th^ March-23^rd^ August 2020). Further, it sought to explore sociodemographic and health-related factors that might be associated with different patterns of longitudinal changes in physical activity. Given the well-established health benefits of physical activity, understanding changes in physical activity habits, and predictors of these changes, is essential for informing healthcare policy in the aftermath of COVID-19.

## Method

### Study design and participants

We analysed data from the COVID-19 Social Study, a longitudinal study run by University College London that focuses on the psychological and social experiences of adults living in the UK during the COVID-19 pandemic. The study commenced on 21^st^ March 2020 and involved weekly online data collection from participants until 23^rd^ August 2020, followed by monthly data collection for the duration of the pandemic. The study design is described in detail elsewhere^22^ and a full protocol is available online (www.COVIDSocialStudy.org).

To examine trajectories of physical activity in relation to specific lockdown measures, we focused solely on participants who lived in England (N=56,428). We included participants who had at least three repeated measures between March 24^th^ 2020, the day after the first lockdown started in the UK, and August 23^rd^ 2020, when the survey switched to monthly follow-up and the relevant measure was discontinued. This period encompasses the first national lockdown followed by the easing of restrictions allowing unlimited outdoor exercises (13 May 2020), reopening outdoor gyms and playgrounds (4 July 2020) and indoor gyms and swimming pools (25 July 2020). These criteria provided us with data from 38,917 participants who were followed up for a maximum of 22 weeks. After excluding participants with missing values (8%), our final analytic sample size was 35,915.

### Measures

In the UK COVID-19 Social Study, participants were asked weekly how long they had spent on the last working day (1) going out for a walk or other gentle physical activity, (2) going out for moderate or high intensity activity (e.g. running, cycling or swimming), (3) exercising inside their home or garden (e.g. doing yoga, weights or other indoor exercise). Responses were recorded on a five-point frequency scale: did not do, <30 minutes, 30 minutes-2 hours, 3-5 hours and ≥6 hours. We generated a composite physical activity measure by using the highest response across these three questions, which was recoded into four categories: (1) did not do, (2) <30 minutes, (3) 30 minutes-2 hours and (4) ≥3 hours. For instance, if a participant walked for ≥3 hours, did high intensity activity for 30 minutes-2 hours, and did not exercise at home, they would be coded as ≥3 hours. As the physical activity questions referred to the last working day, and the first national lockdown started on 23^rd^ March 2020, we included responses from 24^th^ March 2020 onwards.

A range of socio-demographic and health-related factors were considered as potential predictors of physical activity trajectories. These included gender (women, men), ethnicity (white, ethnic minorities), age groups (18-29, 30-45, 46-59, 60+ years), education (GCSEs or below, A-levels or equivalent, degree or above), household income (<£30,000, >£30,000 per annum), employment status (employed throughout, employed at baseline but lost job during the follow-up, unemployed or economically inactive), living arrangement (living alone, living with others but no children, living with others including children), and area of living (city, large town, small town, rural). Health-related factors were self-reported diagnosis of any long-term physical health condition, including disability (yes, no), and self-reported diagnosis of any long-term mental health condition (yes, no).

### Statistical analysis

Data were analysed using the growth mixture modelling (GMM) approach. The conventional growth modelling approach assumes one homogeneous growth trajectory, allowing individual growth factors to vary randomly around the overall mean. GMM relaxes this assumption and enables exploration of different patterns of change (latent trajectory classes).^23^

We included a polynomial time function to allow for nonlinear growth trajectories informed by the data. Starting with the unconditional GMM, we compared models with different number of classes on the basis of the Bayesian information criterion (BIC) and sample-size adjusted Bayesian information criterion (ABIC), along with the Vuong-Lo-Mendell-Rubin likelihood ratio (LMR-LR) test and Adjusted Lo-Mendell-Rubin likelihood ratio (ALMR-LR) test. After identifying the optimal number of classes, we introduced predictors to explain the observed heterogeneity between classes.

Weights were applied throughout the analyses. The final sample was weighted to the proportions of gender, age, ethnicity and education in the English population obtained from the Office for National Statistics.^24^ Analyses were implemented in Mplus Version 8.

## Results

### Descriptive

The analytical sample comprised 35,915 participants, of whom 75.6% were women (Table 1).There was an over-representation of people with a degree or above (70.3%) and an underrepresentation of people from ethinic minority backgrounds (5.0%) and younger adults under 30 (7.4%). After weighting, the sample reflected population proportions (Table 1).

**Table 1.** Characteristics of the sample (N=35,915)

|  | Raw data |  | Weighted data |  |
| --- | --- | --- | --- | --- |
|  | Percent | N | Percent | N |
| **Gender** |  |  |  |  |
| Women | 75.9% | 27,245 | 51.0% | 18,330 |
| Men | 24.1% | 8,670 | 49.0% | 17,585 |
| **Ethnicity** |  |  |  |  |
| Minority | 5.0% | 1,796 | 14.5% | 5,223 |
| White | 95.0% | 34,119 | 85.5% | 30,692 |
| **Age** |  |  |  |  |
| 18-29 | 7.4% | 2,656 | 19.5% | 7,012 |
| 30-45 | 29.0% | 10,431 | 26.4% | 9,483 |
| 46-59 | 33.0% | 11,858 | 24.1% | 8,657 |
| 60+ | 30.5% | 10,970 | 30.0% | 10,763 |
| **Education** |  |  |  |  |
| GCSE or below | 12.9% | 4,636 | 32.5% | 11,656 |
| A-levels or equivalent | 16.8% | 6,031 | 32.9% | 11,807 |
| Degree or above | 70.3% | 25,248 | 34.7% | 12,452 |
| **Household income** |  |  |  |  |
| <30k | 36.7% | 13,193 | 46.0% | 16,519 |
| ≥30k | 63.3% | 22,722 | 54.0% | 19,396 |
| **Employment status** |  |  |  |  |
| Employed | 55.7% | 19,988 | 49.0% | 17,587 |
| Employed to unemployed | 10.4% | 3,724 | 10.1% | 3,636 |
| Unemployed/economically inactive | 34.0% | 12,203 | 40.9% | 14,692 |
| **Living status** |  |  |  |  |
| Alone | 19.8% | 7,100 | 18.3% | 6,577 |
| With others (not children) | 53.2% | 19,106 | 56.2% | 20,168 |
| With others (including children) | 27.0% | 9,709 | 25.5% | 9,169 |
| **Area of living** |  |  |  |  |
| City | 35.4% | 12,717 | 34.8% | 12,503 |
| Large town | 19.1% | 6,859 | 21.8% | 7,817 |
| Small town | 24.8% | 8,916 | 24.4% | 8,770 |
| Rural area | 20.7% | 7,423 | 19.0% | 6,825 |
| **Long term physical illness** |  |  |  |  |
| Yes | 39.2% | 14,086 | 40.8% | 14,651 |
| No | 60.8% | 21,829 | 59.2% | 21,264 |
| **Long term mental illness** |  |  |  |  |
| Yes | 18.3% | 6,578 | 20.0% | 7,186 |
| No | 81.7% | 29,337 | 80.0% | 28,729 |

Figure 1 shows how the proportion of the sample in each physical activity category changed over the course of 22 weeks from the start of lockdown. There is little evidence that the overall prevalence of physical activity showed sudden, concurrent changes with major adjustments in UK lockdown measures.

**Figure 1.**
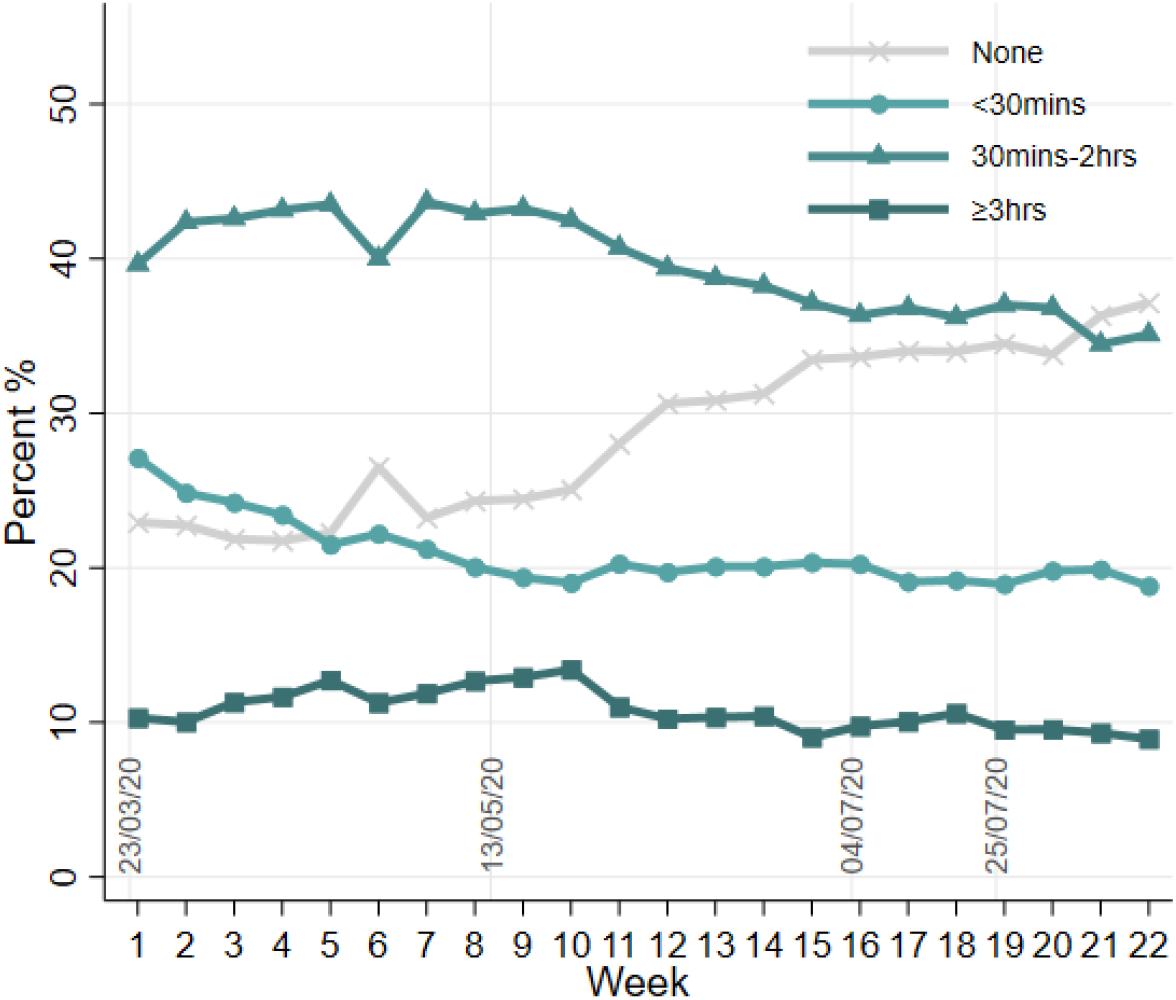
Descriptive changes of physical activity over 22 weeks. On March 23 2020, the first lockdown commenced in England. All non-essential businesses, including gyms, outdoor sports amenities, and playgrounds were closed. On May 10 2020, it was announced that strict lowdown was being eased, with unlimited outdoor exercise being allowed from May 13 2020. On July 4, further public amenities were reopened, including outdoor gyms and playgrounds. On July 25 2020, indoor gyms and swimming pools reopened.

### Latent trajectory classes

To determine the optimal number of latent trajectory classes, we estimated and compared across unconditional GMMs with different numbers of classes. Although the BIC and ABIC decreased with each additional class added to the model, the ALMR-LR test of the seven-class GMM did not reject the six-class model (Table S1). Therefore, the six-class GMM model was chosen. It had an adequate quality of class membership classification (entropy=0.72). The estimated probability of each physical activity category for each latent class (LC) is presented in Figure 2.

**Figure 2.**
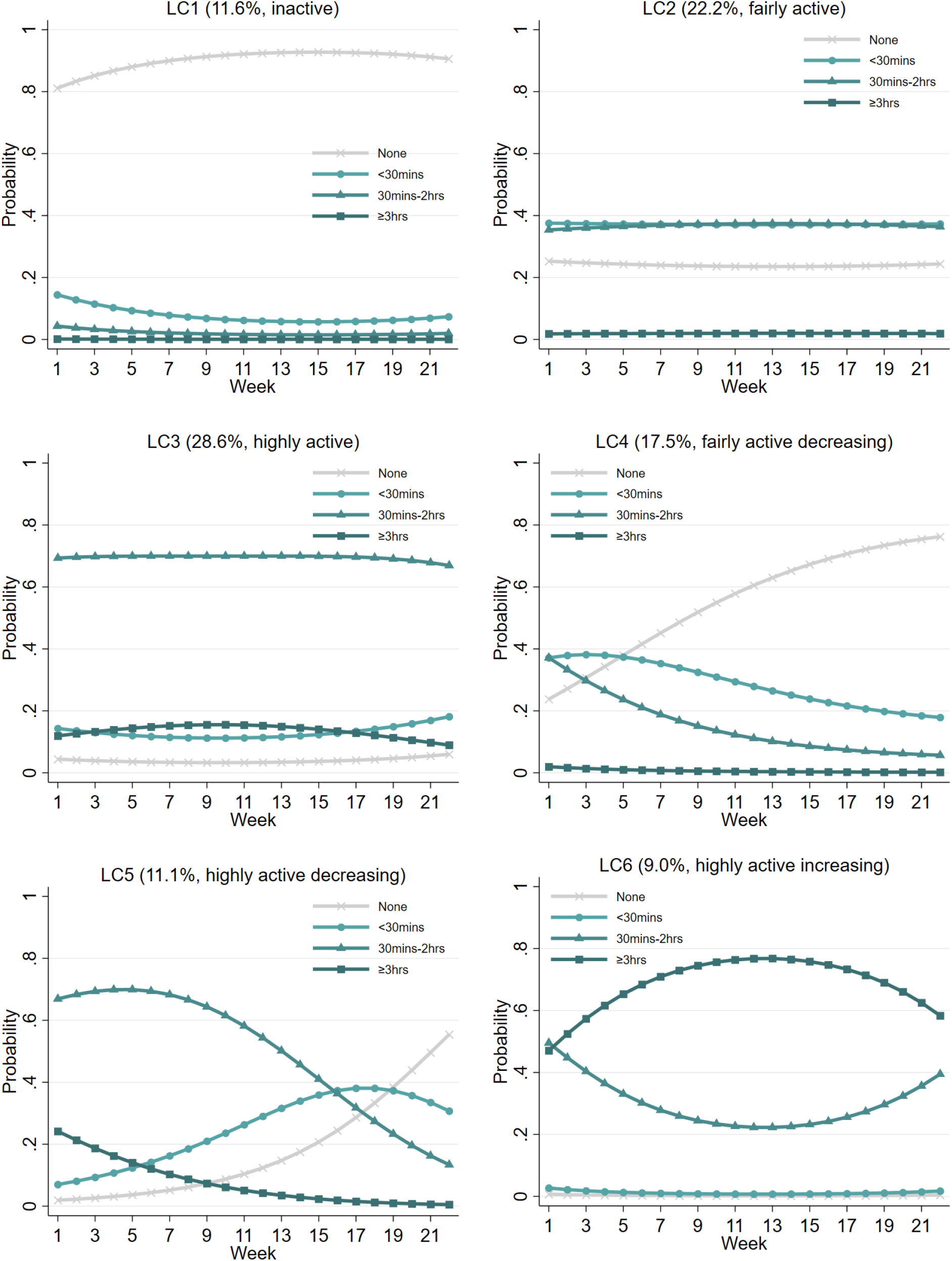
Estimated growth trajectories for different classes.

The first three classes (LC1-3) were static, with little change observed over the 22-week period, making up 62.4% of the sample. The first class (LC1=11.6%) was marked by a high probability of physical inactivity (“inactive”). Participants in the second class (LC2=22.2%) had a moderate probability of doing physical activity for short (under 30 minutes) and medium (30 minutes to 2 hours) durations (“fairly active”). The third class (LC3=28.6%) was the largest class, which consisted of people with a high probability of exercising for 30 minutes to 2 hours (“highly active”).

The last three classes (LC4-6) were dynamic, showing substantial changes over time. Both class four and five, together forming 28.6% of the sample, showed decreasing physical activity over time. The fourth class (LC4=17.5%) started from a moderate level of physical activity, very similar to LC2 when the lockdown started. In this class, the probability of being inactive increased rapidly over time, which was accompanied by declines in the short (<30 minutes) and medium-duration (30 minutes to 2 hours) categories (“fairly active decreasing”). In contrast, participants in LC5 (11.1%) started from a high probability of exercising for 30 minutes to 2 hours, similar to LC3 at the beginning of lockdown. This probability was stable over the first few weeks but was followed by a rapid decline in subsequent weeks (“highly active decreasing”). This decline translated into an increased probability of being in the inactive or short duration (<30 minutes) categories. The sixth class (LC6=9.0%) was the smallest and the most active, showing a high probability of exercising for medium (30 minutes to 2 hours) and long (≥3 hours) durations. It was the only class that showed an overall increase in physical activity over time (“highly active increasing”). More specifically, the probability of exercising for a long duration (≥3 hours) increased in the first 13 weeks, which was followed by a decrease when lockdown measures were substantially eased in June 2020. The growth trajectory of the long duration category (≥3 hours) was the opposite of that for the category of medium duration (30 minutes to 2 hours), indicating exclusive transition between these two categories. Notably, this class had a very low probability of exercising for a short duration (<30 minutes) or being physically inactive, which did not change over time.

### Factors associated with latent trajectory classes

We fitted a conditional GMM to examine how individual characteristics were related to class membership of physical activity trajectories, using LC1 (“inactive”) as the reference (Table 2). Young adults had higher odds of being “fairly active” (LC2) than people aged 30 to 45 (OR=1.48, 95% CI=1.03-2.13), as did individuals with a degree or above compared to those with lower education (OR=2.06, 95% CI=1.66-2.56). People living with children also had higher odds of being “fairly active” than those living alone (OR=1.54, 95% CI=1.18-2.00). However, people with long-term physical (OR=0.83, 95% CI=0.69-1.00) and mental (OR=0.57, 95% CI=0.46-0.71) health conditions had lower odds of being “fairly active”.

**Table 2.** Results from the Growth mixture model with predictors of latent classes (LC) (LC1 as the reference, N=35,915)

|  | LC2 (vs. LC1) |  | LC3 (vs. LC1) |  | LC4 (vs. LC1) |  | LC5 (vs. LC1) |  | LC6 (vs. LC1) |  |
| --- | --- | --- | --- | --- | --- | --- | --- | --- | --- | --- |
|  | OR | 95% CI | OR | 95% CI | OR | 95% CI | OR | 95% CI | OR | 95% CI |
| Woman (vs man) | 1.02 | [0.85-1.23] | 0.96 | [0.81-1.14] | 1.19 | [0.98-1.46] | 1.19 | [0.94-1.49] | **0.71** | **[0.58-0.87]** |
| Ethnic minority (vs white) | 0.82 | [0.57-1.17] | 0.89 | [0.62-1.29] | 0.99 | [0.66-1.48] | 0.83 | [0.52-1.31] | 1.04 | [0.68-1.59] |
| Age 18-29 (vs. 30-45) | **1.48** | **[1.03-2.13]** | 1.24 | [0.85-1.80] | 1.32 | [0.86-2.03] | **2.01** | **[1.33-3.04]** | **1.62** | **[1.02-2.57]** |
| Age 46-59 (vs. 30-45) | 0.83 | [0.67-1.03] | **1.32** | **[1.07-1.63]** | 0.83 | [0.65-1.06] | 0.94 | [0.72-1.22] | 1.19 | [0.89-1.59] |
| Age 60+ (vs. 30-45) | 1.22 | [0.93-1.59] | **2.23** | **[1.72-2.89]** | 1.18 | [0.87-1.60] | **1.75** | **[1.25-2.45]** | **2.07** | **[1.47-2.92]** |
| A-levels or equivalent (vs. GCSEs or below) | 1.09 | [0.87-1.36] | **1.25** | **[1.01-1.54]** | 1.12 | [0.87-1.43] | **1.44** | **[1.09-1.90]** | 1.08 | [0.83-1.41] |
| Degree or above (vs. GCSEs or below) | **2.06** | **[1.66-2.56]** | **2.36** | **[1.93-2.90]** | **1.35** | **[1.06-1.72]** | **1.78** | **[1.34-2.35]** | 1.27 | [0.97-1.68] |
| Household income <30k (vs ≥30k) | 0.82 | [0.67-1.02] | **0.54** | **[0.44-0.65]** | 0.80 | [0.64-1.01] | **0.46** | **[0.36-0.60]** | **0.66** | **[0.52-0.84]** |
| Employed to unemployed (vs. employed) | 1.25 | [0.90-1.73] | **1.68** | **[1.24-2.26]** | **1.52** | **[1.08-2.13]** | **1.79** | **[1.26-2.56]** | **1.77** | **[1.27-2.48]** |
| Unemployed/inactive (vs. employed) | 0.86 | [0.69-1.08] | 1.06 | [0.87-1.29] | **0.77** | **[0.61-0.99]** | **0.70** | **[0.52-0.94]** | 0.94 | [0.72-1.21] |
| Living with others, but no children (vs alone) | 1.20 | [0.98-1.48] | **1.37** | **[1.13-1.67]** | 1.04 | [0.83-1.30] | **1.60** | **[1.21-2.11]** | **1.29** | **[1.01-1.64]** |
| Living with others, including children (vs. alone) | **1.54** | **[1.18-2.00]** | **1.51** | **[1.17-1.95]** | 1.28 | [0.95-1.74] | **1.60** | **[1.13-2.26]** | **1.62** | **[1.17-2.23]** |
| Large town (vs. city) | 0.83 | [0.65-1.04] | 0.87 | [0.69-1.09] | 1.06 | [0.81-1.38] | 0.83 | [0.61-1.13] | 0.91 | [0.67-1.22] |
| Small town (vs. city) | 0.92 | [0.74-1.15] | 1.03 | [0.82-1.29] | 0.93 | [0.71-1.20] | 0.91 | [0.68-1.21] | 1.00 | [0.75-1.33] |
| Rural (vs. city) | 0.87 | [0.67-1.12] | 1.17 | [0.92-1.48] | 1.30 | [0.97-1.74] | 1.32 | [0.97-1.80] | 1.27 | [0.96-1.67] |
| Long-term physical illness (vs. none) | **0.83** | **[0.69-1.00]** | **0.49** | **[0.41-0.58]** | 0.86 | [0.70-1.04] | **0.55** | **[0.44-0.69]** | **0.61** | **[0.49-0.75]** |
| Long-term mental illness (vs. none) | **0.57** | **[0.46-0.71]** | **0.48** | **[0.39-0.60]** | **0.70** | **[0.56-0.88]** | **0.49** | **[0.36-0.66]** | **0.55** | **[0.43-0.72]** |

Note: p<0.05 in bold text. LC1: inactive, LC2: fairly active, LC3: highly active, LC4: fairly active decreasing, LC5: highly active decreasing, LC6: highly active increasing.

Older adults had higher odds of being “highly active” (LC3) than those aged 30 to 45 (OR=1.32-2.23), as did people with higher levels of education (OR=1.25-2.36), those who lost their job (OR=1.68, 95% CI=1.24-2.26), and people living with others (OR=1.37-1.51). People from low-income households (OR=0.54, 95% CI=0.44-0.65), and those with physical (OR=0.49, 95% CI=0.41-0.58) and mental (OR=0.48, 95% CI=0.39-0.60) health conditions had lower odds of being “highly active”.

For the dynamic classes, individuals with a degree or above (OR=1.35, 95% CI=1.06-1.72) and those who lost their job (OR=1.52, 95% CI=1.08-2.13) had higher odds of “fairly active decreasing” activity (LC4). People who were already unemployed or economically inactive at the start of lockdown (OR=0.77, 95% CI=0.61-0.99) and those with mental health conditions (OR=0.70, 95% CI=0.56-0.88) had lower odds of being in LC4.

Young adults (OR=2.01, 95% CI=1.33-3.04) and older adults (OR=1.75, 95% CI=1.25-2.45) had higher odds of “highly active decreasing” activity (LC5) than those aged 30 to 45. Also more likely to be in this class were people who: had higher levels of education (OR=1.44-1.78); lost their job (OR=1.79, 95% CI=1.26-2.56); and were living with others (OR=1.60-1.60). Conversely, people who were from low-income households (OR=0.46, 95% CI=0.36-0.60), unemployed or economically inactive (OR=0.70, 95% CI=0.52-0.94), and had physical (OR=0.55, 95% CI=0.44-0.69) and mental (OR=0.49, 95% CI=0.36-0.66) health conditions had lower odds of being in LC5.

Finally, young adults (OR=1.62. 95% CI=1.02-2.57), older adults (OR=2.07, 95% CI=1.47-2.92), people who lost their job (OR=1.77, 95% CI=1.27-2.48) and those living other others (OR=1.29-1.62) had higher odds of “highly active increasing” activity (LC6). In contrast, women (OR=0.71, 95% CI=0.58-0.87), people from low-income households (OR=0.66, 95% CI=0.52-0.84) and those with physical (OR=0.61, 95% CI=0.49-0.75) and mental (OR=0.55, 95% CI=0.43-0.72) health conditions had lower odds of being in LC6, the only class where physical activity increased over time.

Given the similarities between LC4 (“fairly active decreasing”) and LC2 (“fairly active”), and between LC5 (“highly active decreasing”) and LC3 (“highly active”) at the beginning of the lockdown, we compared factors associated with these trajectories using alternative reference classes (Table S2). Individuals who had a degree or above (vs GCSEs or below), those who were unemployed or economically inactive (vs employed), and people aged 46-59 (vs 30-45) had lower odds of being in a class with decreasing physical activity. In contrast, people living in rural areas (vs cities) and aged 18-29 (vs 30-45) had higher odds of decreasing physical activity throughout the pandemic.

We carried out sensitivity analyses excluding keyworkers (n=8,651) who might have had a different experience during the lockdown due to still being able to go to work (analytical sample N=27,264). The results were materially consistent with the main analysis, returning the same number of classes and very similar patterns of growth trajectories (Figure S1). Other sensitivity analyses using piecewise growth models to reflect changes in lockdown measures also yielded similar results (Figure S2).

## Discussion

This study is the first to examine the heterogeneity in longitudinal changes in physical activity during the COVID-19 pandemic. Building on recent longitudinal studies, which reported a general decline in physical activity at the start of the pandemic,^10–15,25^ our analyses identified six unique classes of growth trajectories of physical activity. Three of these classes were stable, showing little change over time, including the inactive (11.6%), the fairly active (22.2%) and the highly active (28.6%). There were two classes showing declines in physical activity or increased physical inactivity, making up 28.6% of all participants. In contrast, 9% of participants showed an upward trend in physical activity over the observational period. These differing trajectories may explain the inconsistent findings to date from longitudinal studies testing whether physical activity returned to pre-lockdown levels with the easing of restrictions in the UK.^10,11,14,15^

This study further examined sociodemographic and health-related predictors of physical activity growth trajectories. When comparing the three stable classes (inactive, fairly active, and highly active; LC1-3), we found no gender, ethnic, or urban/rural differences between them. However, people who were older, more educated, had a higher income, shared a household with others, and those without long-term physical or mental health problems, were more likely to be in a more active class. This is consistent with previous evidence that age, education, income, health status, and social support are associated with physical activity during lockdown.^11,15,16,20,21^ In contrast, our findings are not consistent with prior evidence that women and people of non-white ethnicity are less active during lockdown, ^12,15,20^ but consistent with a review of reviews suggesting gender and ethnicity are correlates but not determinants of physical activity.^26^ Previous studies have also found that differences in physical activity between genders and ethnic groups are very small.^27^

A similar set of factors were found to predict the difference between the three dynamic classes (LC4-LC6) relative to the inactive (LC1). The dynamic classes were either fairly active or highly active at the start of lockdown, followed by decreases or increases in physical activity during lockdown. As with the stable classes, people who had higher education and income, lived with others, and did not have long-term health problems were more likely to be in the dynamic fairly or highly active classes than the inactive class. Additionally, individuals who became unemployed were more likely to be in the highly active or dynamic classes. It is possible that this group was unique in having to adjust how they spent their time during lockdown after becoming unemployed, compared to those who were employed or economically inactive throughout this period. Further research with this group may enable us to identify opportunities to increase physical activity and understand barriers that prevent this behaviour change.

There were very few sociodemographic or health-related differences in the dynamic compared to stable classes that started at similar levels of physical activity (fairly active decreasing vs fairly active, and highly active decreasing vs highly active). However, those with decreasing levels of physical activity (i.e. in dynamic classes) were younger, more likely to be employed, living in rural areas, and had lower levels of education. Understanding why these factors were associated with decreasing physical activity is important for the development of interventions. Although younger people were generally more physically active before the pandemic,^26^ several other studies have found that younger adults were more likely to report changes (both increases and decreases) in physical activity during lockdown than older adults.^20,28^ This could be because younger adults were generally confined to smaller homes with no outdoor space and less space to exercise, meaning motivation to remain physically active reduced over time. Levels of physical activity in younger adults may also have decreased as restrictions eased and they replaced time previously spent on physical activity with more sedentary leisure activities, such as socialising. Additionally, being younger and having lower educational attainment are associated with higher levels of anxiety, depression, and loneliness during lockdown,^22,29^ all of which could have contributed to reductions in physical activity. It is also possible that decreasing physical activity was associated with being employed because individuals struggled to maintain levels of physical activity alongside working from home, decreased work-life balance, and increased stress and burnout throughout the lockdown.^30^

This study has a number of strengths including its large sample size, repeated weekly follow-up of the same participants over 22 weeks since the first UK lockdown, and robust statistical approaches. Although the UCL COVID-19 Social Study did not use a random sample, it does have a large sample size, including good stratification across all major socio-demographic groups. In addition, analyses were weighted on the basis of population estimates of core demographics, with the weighted data showing good alignment with national population statistics and another nationally representative social survey.^29^ Despite efforts to make our sample inclusive and representative of the adult population in England, we cannot rule out potential biases due to the omission of other demographic factors, which could be associated with survey participation, from the weighting process. Further, our analyses relied on self-reported time spent on physical activity which is subject to recall and reporting bias. We also lack data on people’s physical activity before the start of the COVID-19 pandemic, so how physical activity levels observed in this study compare to pre-pandemic physical activity in this cohort remains unknown. Finally, this study only includes data from during and shortly after the first UK lockdown. Future studies could extend our analyses to explore whether, and to what extent, the longitudinal patterns of physical activity persist after the COVID-19 pandemic.

Although there may have been a decline in physical activity in the general population at the start of the COVID-19 pandemic, it is important to acknowledge the heterogeneity in people’s longitudinal changes in physical activity. We have shown that over 62% of people experienced little change and another 9% increased their physical activity between March and August 2020. However, it should be highlighted that nearly 29% of people experienced reduced physical activity during the same period. Moreover, amongst the people with little change in physical activity, 12% were consistently inactive. Both of these groups call for attention and action. Given the well-established link between physical activity and health,^1–3^ persistent or increased physical inactivity is likely to have both immediate and long-term implications for people’s physical and mental health, as well as general wellbeing. More public health efforts should be made to promote physical activity for the general population, and in particular for groups that are at a higher risk of inactivity or of reduced physical activity, during the COVID-19 pandemic and beyond.

## Data Availability

Anonymous data will be made publicly available following the end of the pandemic.

## Declarations

### Ethics approval and consent to participate

The study was approved by the UCL Research Ethics Committee [12467/005] and all participants gave informed consent.

### Data availability

Anonymous data will be made publicly available following the end of the pandemic.

### Author contributions

FB, AS and DF developed the study idea and the analysis plan. FB analysed the data. JB and JM did the literature review. FB, JB and JM wrote the first draft. FB, JB and JM had accessed and verified the underlying data. All authors provided critical revisions. All authors read and approved the submitted manuscript.

## Supplementary Material

**Table S1.** Model fit indices for different model specifications

| Model specification | BIC | ABIC | LMR-LR | ALMR-LR | Entropy |
| --- | --- | --- | --- | --- | --- |
| 1-class GMM | 1,098,424 | 1,098,408 | NA | NA | NA |
| 2-class GMM | 947,350 | 947,322 | <0.001 | <0.001 | 0.855 |
| 3-class GMM | 903,370 | 903,329 | <0.001 | <0.001 | 0.857 |
| 4-class GMM | 874,221 | 874,167 | <0.001 | <0.001 | 0.814 |
| 5-class GMM | 867,525 | 867,459 | <0.001 | <0.001 | 0.765 |
| **6-class GMM** | **862,970** | **862,890** | **<0.001** | **<0.001** | **0.723** |
| 7-class GMM | 860,626 | 860,534 | 0.199 | 0.209 | 0.728 |

Notes: Models with smaller BIC and ABIC have a better fit. LMR-LR and ALMR-LR compare model fit between models with k classes and (k – 1) classes. A significant p value indicates a significant model fit improvement in the k-class model. Entropy is a measure of the quality of class membership classification. A value of 0.80 is considered as high, 0.60 is medium, and 0.40 is low

**Table S2.** Results from the Growth mixture model with predictors of latent classes (LC) using alternative reference class (N=35,915)

|  | LC4 (vs. LC2) |  | LC5 (vs. LC3) |  | LC5 (vs. LC4) |  | LC6 (vs. LC3) |  |
| --- | --- | --- | --- | --- | --- | --- | --- | --- |
|  | OR | 95% CI | OR | 95% CI | OR | 95% CI | OR | 95% CI |
| Woman (vs man) | 1.17 | [0.96-1.41] | 1.24 | [1.01-1.52] | 0.99 | [0.80-1.24] | **0.74** | **[0.62-0.88]** |
| Ethnic minority (vs white) | 1.21 | [0.81-1.80] | 0.92 | [0.58-1.47] | 0.84 | [0.54-1.30] | 1.16 | [0.78-1.73] |
| Age 18-29 (vs. 30-45) | 0.89 | [0.61-1.29] | **1.63** | **[1.10-2.39]** | **1.52** | **[1.06-2.19]** | 1.31 | [0.87-1.97] |
| Age 46-59 (vs. 30-45) | 1.00 | [0.79-1.25] | **0.71** | **[0.56-0.91]** | 1.14 | [0.89-1.46] | 0.90 | [0.69-1.17] |
| Age 60+ (vs. 30-45) | 0.97 | [0.74-1.28] | 0.79 | [0.58-1.07] | **1.48** | **[1.07-2.04]** | 0.93 | [0.69-1.26] |
| Employed to unemployed (vs. employed) | 1.03 | [0.80-1.32] | 1.15 | [0.88-1.51] | 1.29 | [0.99-1.68] | 0.86 | [0.68-1.10] |
| Unemployed/inactive (vs. employed) | **0.65** | **[0.52-0.82]** | **0.75** | **[0.57-0.98]** | 1.32 | [1.00-1.74] | **0.54** | **[0.42-0.69]** |
| Household income <30k (vs ≥30k) | 0.97 | [0.79-1.20] | 0.86 | [0.68-1.08] | **0.57** | **[0.45-0.73]** | 1.23 | [1.00-1.52] |
| Employed to unemployed (vs. employed) | 1.21 | [0.89-1.66] | 1.07 | [0.79-1.44] | 1.18 | [0.87-1.60] | 1.06 | [0.81-1.38] |
| Unemployed/inactive (vs. employed) | 0.89 | [0.70-1.15] | **0.66** | **[0.50-0.87]** | 0.90 | [0.67-1.22] | 0.88 | [0.70-1.11] |
| Living with others, but no children (vs alone) | 0.86 | [0.69-1.07] | 1.17 | [0.89-1.53] | **1.54** | **[1.17-2.03]** | 0.94 | [0.75-1.16] |
| Living with others, including children (vs. alone) | 0.84 | [0.64-1.10] | 1.06 | [0.76-1.47] | 1.25 | [0.90-1.73] | 1.07 | [0.81-1.42] |
| Large town (vs. city) | 1.28 | [0.99-1.65] | 0.96 | [0.71-1.28] | 0.79 | [0.58-1.06] | 1.04 | [0.80-1.37] |
| Small town (vs. city) | 1.01 | [0.80-1.26] | 0.88 | [0.67-1.16] | 0.98 | [0.74-1.29] | 0.97 | [0.76-1.25] |
| Rural (vs. city) | **1.50** | **[1.15-1.95]** | 1.13 | [0.86-1.49] | 1.02 | [0.76-1.37] | 1.08 | [0.86-1.36] |
| Long-term physical illness (vs. none) | 1.03 | [0.85-1.25] | 1.12 | [0.90-1.39] | **0.64** | **[0.52-0.80]** | **1.24** | **[1.02-1.50]** |
| Long-term mental illness (vs. none) | 1.23 | [0.97-1.56] | 1.01 | [0.72-1.42] | **0.69** | **[0.51-0.94]** | 1.15 | [0.88-1.50] |

Note: p<0.05 in bold text

**Figure S1.**
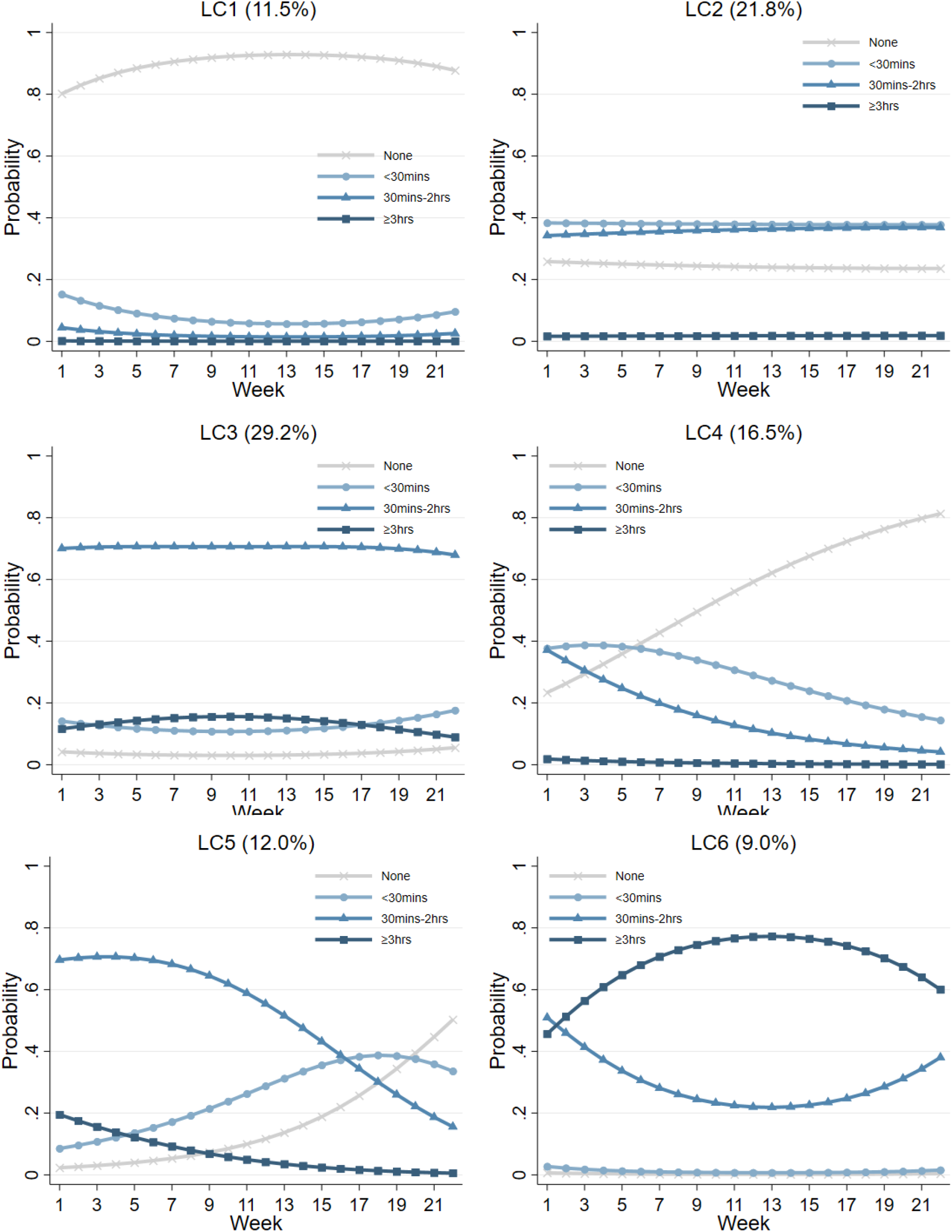
Estimated growth trajectories for different classes (excluding keyworkers)

**Figure S2.**
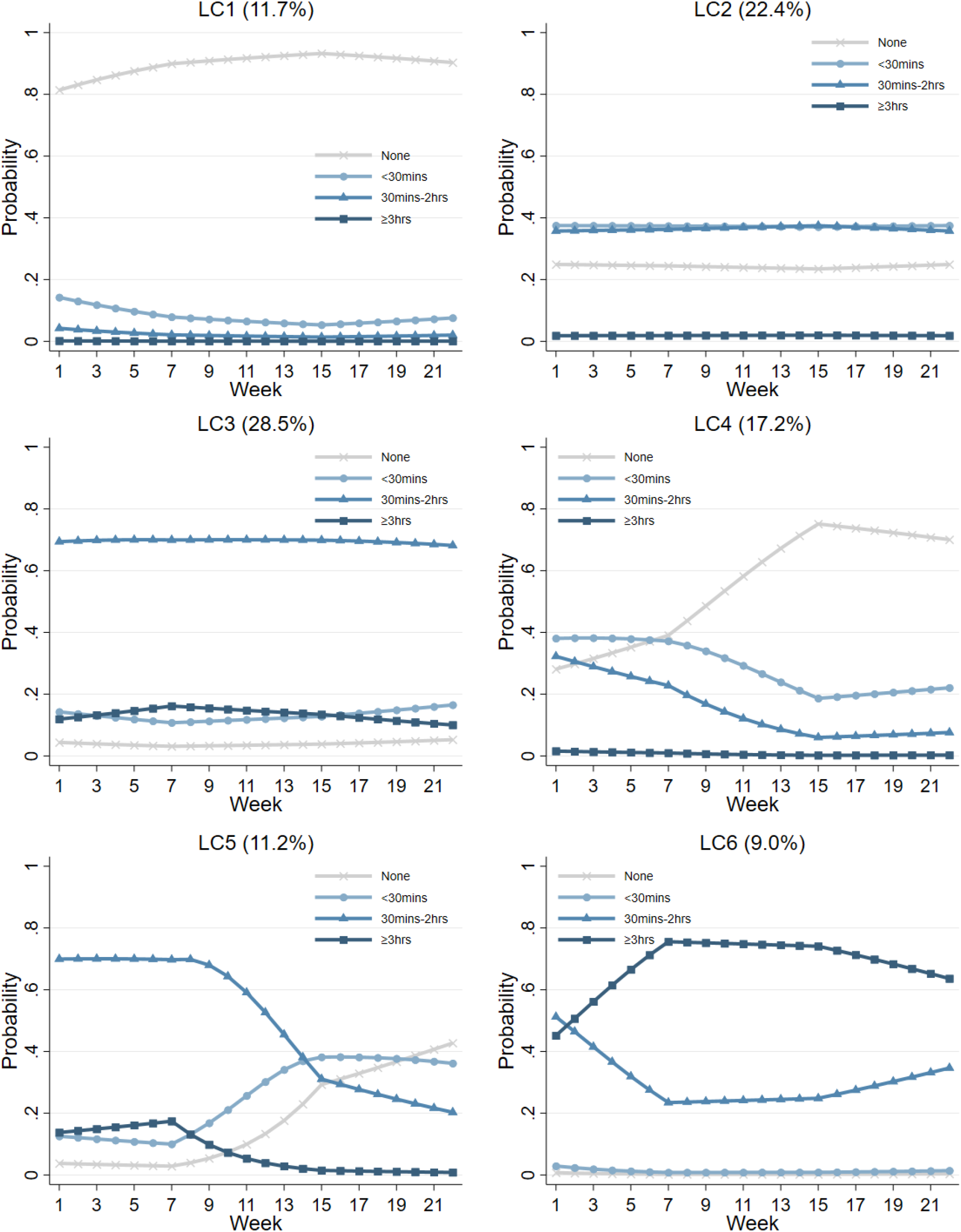
Estimated growth trajectories for different classes (piecewise growth)

## References

1. Penedo, F. J. & Dahn, J. R. Exercise and well-being: A review of mental and physical health benefits associated with physical activity. Curr. Opin. Psychiatry 18, 189–193 (2005).

2. Saint-Maurice, P. F. et al. Association of Daily Step Count and Step Intensity with Mortality among US Adults. JAMA 323, 1151–1160 (2020).

3. World Health Organization. Global recommmendations on physical activity for health. (2010).

4. Carter, S., Hartman, Y., Holder, S., Thijssen, D. H. & Hopkins, N. D. Sedentary behavior and cardiovascular disease risk: Mediating mechanisms. Exerc. Sport Sci. Rev. 45, 80–86 (2017).

5. Nosova, E. V. et al. Short-term physical inactivity impairs vascular function. J. Surg. Res. 190, 672–682 (2014).

6. Bourdas, D. I. & Zacharakis, E. D. Evolution of changes in physical activity over lockdown time: Physical activity datasets of four independent adult sample groups corresponding to each of the last four of the six COVID-19 lockdown weeks in Greece. Data Br. 32, 106301 (2020).

7. Faulkner, J. et al. Physical activity, mental health and well-being of adults during initial COVID-19 containment strategies: A multi-country cross-sectional analysis. J. Sci. Med. Sport (2020) doi:10.1016/j.jsams.2020.11.016.

8. Violant-Holz, V. et al. Psychological health and physical activity levels during the covid-19 pandemic: A systematic review. Int. J. Environ. Res. Public Health 17, 1–19 (2020).

9. Ðogaš, Z. et al. The effect of COVID-19 lockdown on lifestyle and mood in Croatian general population: A cross-sectional study. Croat. Med. J. 61, 309–318 (2020).

10. Janssen, X. et al. Changes in physical activity, sitting and sleep across the COVID-19 national lockdown period in Scotland. medRxiv (2020) doi:10.1101/2020.11.05.20226381.

11. McCarthy, H., Potts, H. & Fisher, A. Physical Activity Behaviour Before, During and After COVID-19 Restrictions: Longitudinal Smartphone-Tracking Study of Adults in the United Kingdom. J. Med. Internet Res. 23, e23701 (2021).

12. Wang, Y. et al. Physical Distancing Measures and Walking Activity in Middle-aged and Older Residents in Changsha, China, during the COVID-19 Epidemic Period: Longitudinal Observational Study. J. Med. Internet Res. 22, (2020).

13. Maltagliati, S. et al. Evolution of Physical Activity Habits After a Context Change: the Case of COVID-19 Lockdown. Sport Rxiv preprint, (2020).

14. Tison, G. H. et al. Worldwide Effect of COVID-19 on Physical Activity: A Descriptive Study. Ann. Intern. Med. 173, 767–770 (2020).

15. Mata, J. et al. Health behaviors and mental health during the COVID-19 pandemic: A longitudinal population-based survey. PsyArXiv (2020).

16. Ong, J. L. et al. COVID-19 Related Mobility Reduction: Heterogenous Effects on Sleep and Physical Activity Rhythms. Sleep 44, zsaa179 (2021).

17. FitBit Staff. The Impact Of Coronavirus On Global Activity. (2020).

18. Garmin. 2020: How Garmin Users Prioritized Movement in a Global Pandemic. (2021).

19. Ding, D., Del Pozo Cruz, B., Green, M. A. & Bauman, A. E. Is the COVID-19 lockdown nudging people to be more active: A big data analysis. Br. J. Sports Med. 54, 2019–2020 (2020).

20. Rogers, N. T. et al. Behavioral Change Towards Reduced Intensity Physical Activity Is Disproportionately Prevalent Among Adults With Serious Health Issues or Self-Perception of High Risk During the UK COVID-19 Lockdown. Front. Public Heal. 8, 1–12 (2020).

21. Meyer, J. et al. Changes in physical activity and sedentary behavior in response to covid-19 and their associations with mental health in 3052 us adults. Int. J. Environ. Res. Public Health 17, 1–13 (2020).

22. Fancourt, D., Steptoe, A. & Bu, F. Trajectories of anxiety and depressive symptoms during enforced isolation due to COVID-19 in England: a longitudinal observational study. The Lancet Psychiatry 8, 141–149 (2021).

23. Muthén, B. & Asparouhov, T. Growth mixture modeling: Analysis with non-Gaussian random effects. in Longitudinal data analysis (eds. Fitzmaurice, G., Davidian, M., Verbeke, G. & Molenberghs, G.) 143–165 (CRC Press, 2008).

24. Knell, G., Robertson, M. C., Dooley, E. E., Burford, K. & Mendez, K. S. Health behavior changes during covid-19 pandemic and subsequent “stay-at-home” orders. Int. J. Environ. Res. Public Health 17, 6268 (2020).

25. Balanzá-Martínez, V. et al. The assessment of lifestyle changes during the COVID-19 pandemic using a multidimensional scale. Rev. Psiquiatr. Salud Ment. 14, 16–26 (2021).

26. Bauman, A. E. et al. Correlates of physical activity: Why are some people physically active and others not? Lancet 380, 258–271 (2012).

27. Rhodes, R. E., Lubans, D. R., Karunamuni, N., Kennedy, S. & Plotnikoff, R. Factors associated with participation in resistance training: A systematic review. Br. J. Sports Med. 51, 1466– 1472 (2017).

28. Rhodes, R. E., Liu, S., Lithopoulos, A., Zhang, C. Q. & Garcia-Barrera, M. A. Correlates of Perceived Physical Activity Transitions during the COVID-19 Pandemic among Canadian Adults. Appl. Psychol. Heal. Well-Being 12, 1157–1182 (2020).

29. Bu, F., Steptoe, A. & Fancourt, D. Who is lonely in lockdown? Cross-cohort analyses of predictors of loneliness before and during the COVID-19 pandemic. Public Health 186, 31–34 (2020).

30. Anderson, D. & Kelliher, C. Enforced remote working and the work-life interface during lockdown. Gend. Manag. 35, 677–683 (2020).

